# Data From the COVID-19 Epidemic in Florida Suggest That Younger Cohorts Have Been Transmitting Their Infections to Less Socially Mobile Older Adults

**DOI:** 10.1101/2020.06.30.20143842

**Authors:** Jeffrey E. Harris

## Abstract

We analyzed the daily incidence of newly reported COVID-19 cases among adults aged 20-39 years, 40-59 years, and 60 or more years in the sixteen most populous counties of the state of Florida from March 1 through June 27, 2020. In all 16 counties, an increase in reported COVID-19 case incidence was observed in all three age groups soon after the governor-ordered Full Phase 1 reopening went into effect. Trends in social mobility, but not trends in testing, track case incidence. Data on hospitalization do not support the hypothesis that the observed increase in case incidence was merely the result of liberalization of testing criteria. Parameter estimates from a parsimonious two-group heterogeneous SIR model strongly support the hypothesis that younger persons, having first acquired their infections through increasing social contact with their peers, then transmitted their infections to older, less socially mobile individuals. Without such cross-infection, an isolated epidemic among older people in Florida would be unsustainable.

---

This study relies exclusively on publicly available, aggregate health data that contain no individual identifiers. The author has no competing interests and no funding sources to declare. This article represents to the sole opinion of its author and does not necessarily represent the opinions of the Massachusetts Institute of Technology, the National Bureau of Economic Research, Eisner Health, or any other organization.

## Introduction

Recent reports suggest that the age distribution of new cases of COVID-19 in the United States has shifted toward younger adults (Malmgren, Guo, and Kaplan 2020). One possible explanation is that younger adults have tended to adhere less strictly to recommended social distancing measures, especially as many states, counties and municipalities have begun to reopen. A particular concern is that the higher prevalence of active infection among younger individuals will ultimately result in a higher rate of cross-infection in older persons.

In this article, we use publicly available data on confirmed individual COVID-19 cases compiled by the state of Florida to test whether the incidence of new coronavirus infections has in fact been rising more rapidly among younger cohorts. We then explore whether the available data can be used to assess whether cross-infection of older cohorts is already occurring.

To that end, we analyze the daily incidence of newly reported COVID-19 cases among adults aged 20-39 years, 40-59 years, and 60 or more years in the sixteen most populous counties of the state of Florida from March 1 through June 27, 2020. In all 16 counties, we observe an increase in reported COVID-19 case incidence in all three age groups soon after the governor-ordered Full Phase 1 reopening went into effect. Trends in social mobility, but not trends in testing, track case incidence. We then use our data on COVID-19 case incidence to estimate a parsimonious two-group heterogeneous SIR model of the epidemic in each county. Our parameter estimates support the hypothesis that younger persons, having first acquired their infections through increasing social contact with their peers, then transmitted their infections to older, less socially mobile individuals. Without such cross-infection, an isolated epidemic among older people in Florida would be unsustainable.

## Data

### Analytic Sample

We downloaded a data file of confirmed individual COVID-19 cases on June 28, 2020 from the website of the Florida Department of Public Health (Florida Department of Public Health 2020b). The database covered 141,040 cases diagnosed through June 27, 2020, showing the age of the individual, the date of diagnosis, county of residence, and whether hospitalized. We excluded 225 cases listed as diagnosed before March 1, 2020, as well as 222 cases of individuals with unknown age. Focusing on adults, we further excluded 12,572 cases with recorded age less than 20 years, leaving 128,021 cases.

We classified the remaining cases into three age groups: 20–39 years old (“younger”), 40–59 years old (“middle aged”), and 60 years or more (“older”). We further focused on cases among residents of the 16 counties with the largest projected population aged 20 years or more (Population Studies Program 2019). The breakdown by county and age group was as follows:

|  | *20–39 Years* | *40–59 Years* | *60+ Years* | *All Ages* |
| --- | --- | --- | --- | --- |
| *Populous Counties* | 46,293 | 35,112 | 25,564 | 106,969 |
| *Other Counties* | 9,089 | 6,631 | 5,332 | 21,052 |
| *Entire State* | 55,382 | 41,743 | 30,896 | 128,021 |

Our *analytic sample* of 106,969 cases, based on the 16 most populous counties, thus constituted 83.6 percent of all statewide confirmed COVID-19 cases among adults diagnosed through our June 27 *closing date*. Figure 1 shows a map of the sixteen counties in our analytic sample.

**Figure 1.**
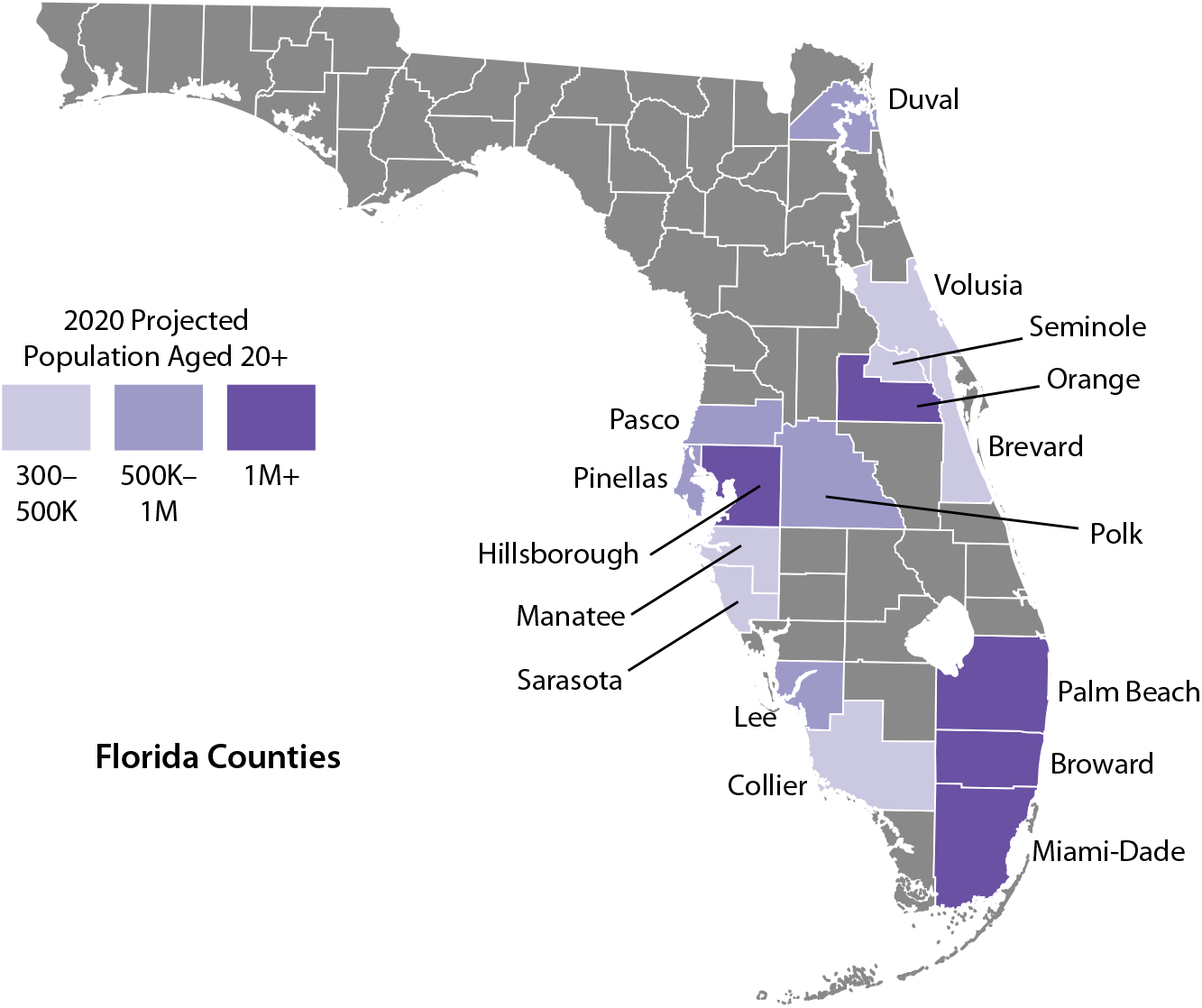
Sixteen Counties in Analytic Sample

### Testing Data

To analyze COVID-19 testing patterns, we further relied on a series of county reports, issued daily by the Florida Department of Public Health from May 13, 2020 through our closing date (Florida Department of Public Health 2020a). We also downloaded data on the daily numbers of positive and negative tests for the entire state of Florida up to the closing date from the COVID Tracking Project website (COVID Tracking Project 2020).

### Social Mobility Data

We accessed two sources of social mobility data: Google’s Community Mobility Reports, which provided information on daily visits to retail establishments and recreational activity for Florida counties (Google 2020); and OpenTable’s data on the daily numbers of seated diners in restaurants in Fort Lauderdale, Miami, Miami Beach, Naples, Orlando, and Tampa (OpenTable 2020).

## Methods

### Descriptive Analyses

We used data on confirmed COVID-19 cases to analyze trends in daily incidence by age group in each of the 16 most populous counties. We related these trends to key statewide regulatory events. We then analyzed trends in social mobility, trends in testing at both the state and county level, and trends in hospitalization rates of older people to assess whether the liberalization of testing criteria was a factor in determining COVID-19 case incidence.

### Two-Group SIR Model

To quantitatively test the hypothesis that younger cohorts of infected individuals have been cross-infecting older persons, we relied on a parsimonious two-group, heterogeneous SIR model (Ellison 2020). To that end, we collapsed the daily COVID-19 incidence data for the younger and middle-aged groups into a single group, ages 20–59 years, retaining the incidence data for the older group, ages 60 years or more.

While we estimated the parameters of our two-group SIR model in discrete time, for clarity of exposition we describe the model here in continuous time. Let 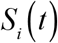, 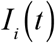, and 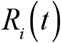 denote the respective proportions of susceptible, infective, and resistant individuals in groups *i =* 1,2 at time *t* ≥ 0. Each group has a closed population, that is, *S_i_* + *I_i_* + *R_i_* = 1 for *i* = 1,2, where we drop the explicit time dependence to simplify the notation. The motion of these state variables over time is governed by four differential equations: 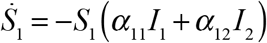, 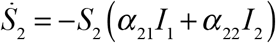, 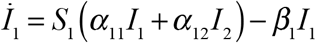, and 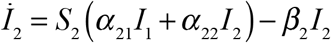, where we have used the notation 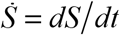 for the time derivative. Given the closed population constraints, the corresponding differential equations for the numbers of resistant individuals are 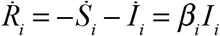 for *i* = 1,2.

The path of this dynamic system is determined by the four positive transmission parameters 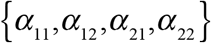, the positive rates at which the two groups become resistant 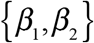, and the initial conditions on the state variables. The transmission parameter *α*_11_ captures the rate at which younger persons come into contact with each other, as well as the probability of transmission when a susceptible younger person comes into contact with an infective younger person. The corresponding parameter *α*_22_ captures the rate at which older persons come into contact with each other, as well as the probability of transmission when a susceptible older person comes into contact with an infective older person. We call these *intragroup* transmission parameters.

Similarly, the *intergroup* transmission parameter *α*_12_ captures the rate at which younger persons come into contact with older persons, as well as the probability of transmission when a susceptible younger person comes into contact with an infective older person. Similarly, the intergroup transmission parameter *α*_21_ captures the rate at which older persons come into contact with younger persons, as well as the probability of transmission when a susceptible older person comes into contact with an infective younger person. The main objective of our modeling effort is to assess the relative magnitude of the latter intergroup transmission parameter *α*_21_.

The parameters 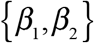 capture the rates at which infective individuals transition to their respective resistant states through recovery or death. While older persons are known to have a significantly higher case mortality rate, we take both parameters to be well approximated by the inverse of the serial interval between successive infections. A recent review gives a range of serial intervals from 3.1 and 7.5 days (Griffin et al. 2020). In our base-case analysis, we assume *β*_1_ = *β*_2_ = *b* = 1/5.5, but vary this parameter from *b* = l/5 to *b* = l/6.

Converting our dynamic model into discrete time, we let *S_ikt_* and *I_ikt_*, respectively, denote the proportions of susceptible and infective individuals in age group *i* = 1,2 located in county *k =* 1,…,16 at date *t*. Let 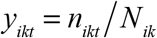 denote the observed incidence rate, which we calculate as the number *n_ikt_* of infections diagnosed among age group *i* in county *k* on date *t*, as derived from our analytic database, divided by *N_ik_*, the projected population of age group *i* in county *k* (Population Studies Program 2019). We then reconstruct the daily path of the epidemic from March 1 through the closing date June 27, 2020 (that is, from *t =* 1,…,119) by iterative application of the discrete equations of motion *S_ikt_* = *S_ik,t_*_−1_ − *y_ikt_* and 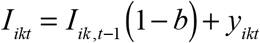, where we take as initial conditions *S_ik_*_0_ = 1 − *I_ik_*_0_ and 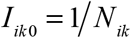. We note that each reconstructed state variable *S_ik,t_*_−1_ and *I_ik,t_*_−1_ depends on the past values of 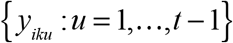 but not on the contemporaneous incidence rate *y_ijt_*.

Our reconstructed epidemic paths in each age group and each county thus provide an empirical framework for estimating the model’s transmission parameters. For notational purposes only, let *X_ijk,t_*_−1_ = *S_ik,t_*_−l_*I_jk,t_*_−l_, where *i* = 1,2 and *j* = 1,2. Then our dynamic equations for the incidence of new daily infections becomes *y_ikt_* = *α_i_*_1_*,X_i_*_1_*_k,t_*_−1_ + *α_i_*_2_*,X_i_*_2_*_k,t_*_−1_ + *ε_ikt_*, where *ε_ikt_* are assumed to be independently identically distributed random variables with zero expectation and, moreover, *ε_ikt_* are independent of *X_ijk,t_*_−l_. In what follows, we focus on the estimation equation for the daily incidence of new infections in older persons, that is *y*_2_*_kt_* = *α*_21_*X*_21_*_k,t_*_−1_ + *α*_22_*X*_22_*_k,t_*_−1_ + *ε*_2_*_kt_*. This equation can be estimated separately for each county *k* or pooled across counties.

## Results

### Daily Incidence by Age Group: 16 Populous Counties Combined

For each adult age group, Figure 2 shows the daily incidence of COVID-19 cases per 100,000 population from March 1 through June 27 for the 16 populous counties combined. The vertical axis is measured on a logarithmic scale so that an exponential rise in incidence would correspond to a straight line on the plot (Harris 2020a). The sky-blue datapoints correspond to the younger age group (20–39 years), the lime datapoints correspond to the middle-aged group (40–59 years), and the mango datapoints represent the older group (60+ years).

**Figure 2.**
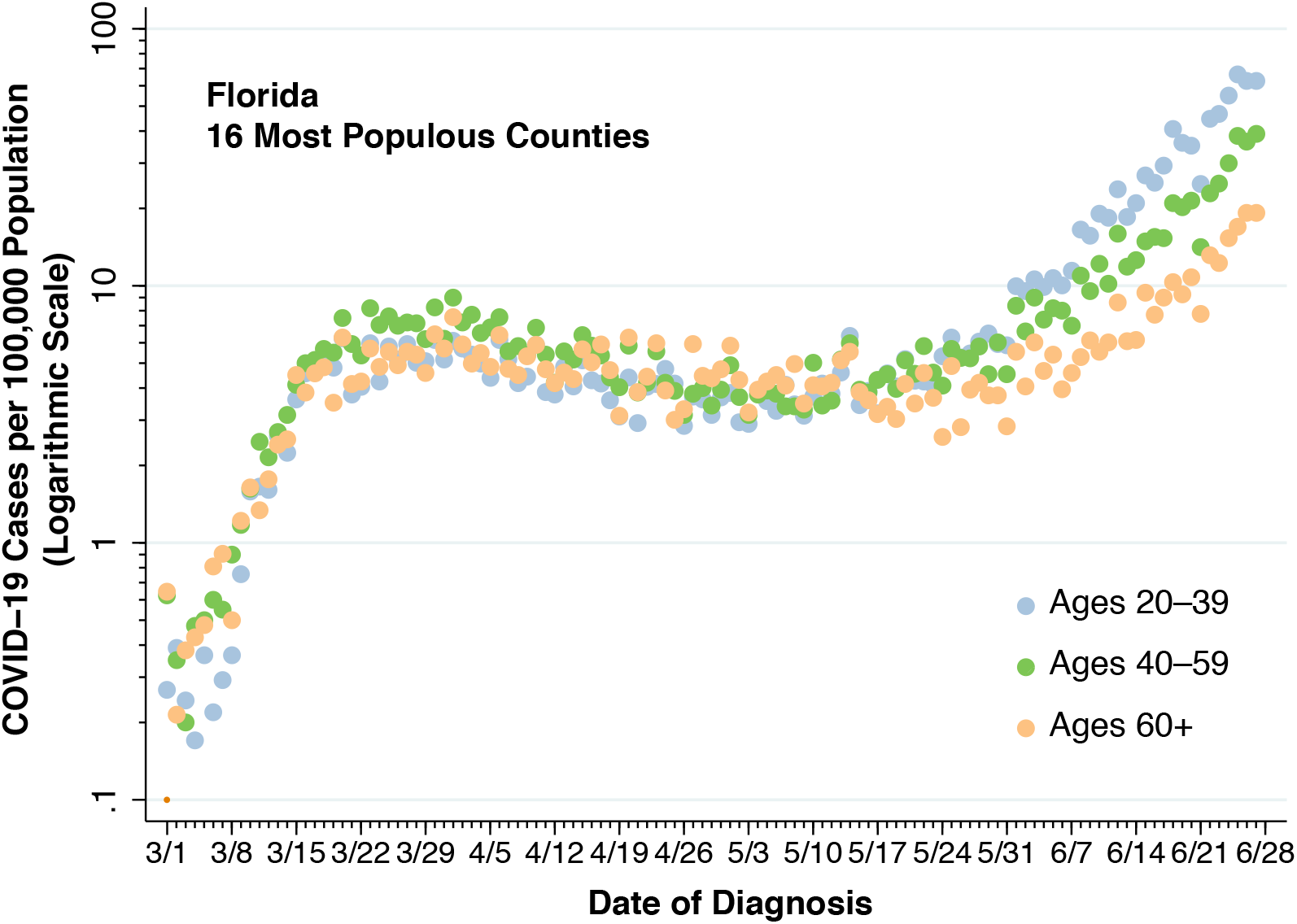
Daily Incidence of New COVID-19 Cases per 100,000 Population by Age Group in the 16 Florida Counties in the Analytic Sample

Figure 2 shows an initial exponential rise in incidence in all three adult age groups during March, followed by a flattening and decline in the incidence curve beginning around Sunday, March 22 and extending to around Sunday, May 17. Since then, the incidence appears to be increasing in all three adult age groups, most markedly in the younger age group (20–39 years).

### Relation Between Trends in Incidence and Key Statewide Regulatory Events

Figure 3 interprets the data displayed in Figure 2. For each calendar week and each age group, we have superimposed the geometric mean incidence,^*^ represented as larger data points connected by line segments. The youngest group corresponds to the larger blue points, the middle-aged group corresponds to the larger green points, and the older group is represented by the larger orange points.

**Figure 3.**
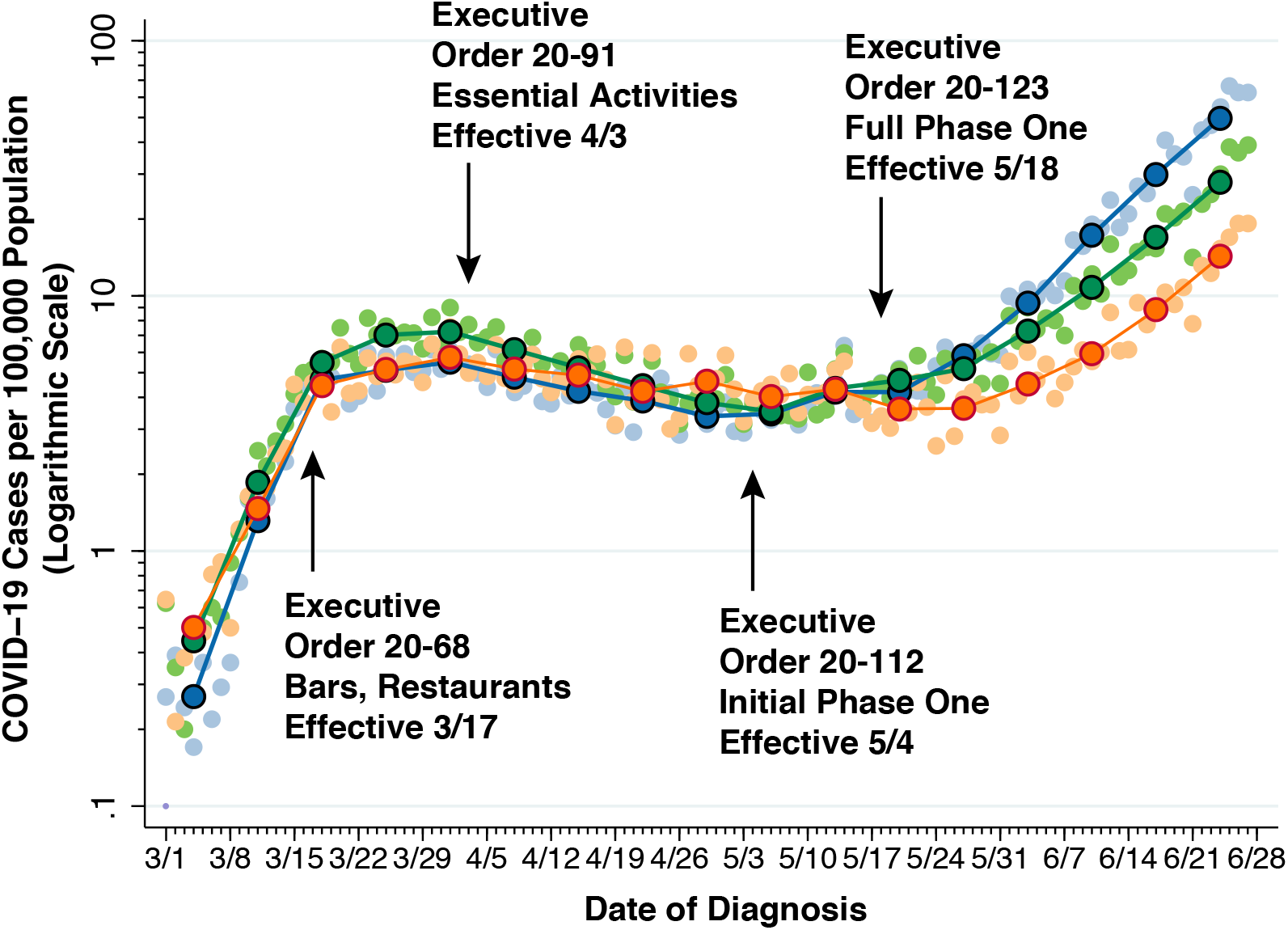
Geometric Mean Daily Incidence Rates by Week and Key Executive Orders Superimposed on the Incidence Plot of Figure 2

The superimposition of the larger connected points helps us see that the incidence has recently been increasing in all three age groups. From the week beginning Sunday May 17 to the week beginning Sunday June 24, the average daily incidence of new COVID-19 cases has increased by 11.83-fold among the younger group, 5.98-fold among the middle-aged group, and 3.96-fold among the older group.

Further superimposed on the incidence trends in Figure 3 are black arrows marking the dates of key orders issued by Florida Gov. Ron DeSantis. Specifically, Order Number 20-68, effective March 17, 2020, imposed restrictions on pubs, bars, nightclubs, restaurants and beaches (Desantis 2020a). Subsequent Order Number 20-91, effective April 3, 2020, limited movement outside the home to essential activities and confined business activities to essential services (Desantis 2020b). Order Number 20-112, effective May 4, 2020, began Phase 1 of the state’s reopening, permitting restaurants and retail stores to operate at 25 percent capacity and liberalizing prior prohibitions on elective medical procedures (Desantis 2020c). Order Number 20-123, effective May 18, 2020, put Full Phase 1 into effect, allowing restaurants, retail establishments, and gyms to operate at 50 percent capacity, opening professional sports events and training camps, and permitting amusement parks and vacation rentals to operate subject to prior approval (Desantis 2020d).

Causal inferences relating trends in COVID-19 incidence to specific regulatory actions must be approached with care (Harris 2020b, c). Still, it is noteworthy that the deceleration of initial exponential surge in early March began soon after Order Number 20-68, while the backward bending of the incidence curve began soon after Order Number 20-91. Moreover, the downward trend in incidence halted soon after Executive Order 20-112, while the resumption of the upward epidemic curve began soon after Executive Order 20-123.

### Trends in Social Mobility

Figure 4 shows the corresponding daily COVID-19 case incidence for the three age groups in Broward County, which includes the city of Fort Lauderdale. As in Figure 3, the incidence is measured in daily cases per 100,000 population for each age group, as measured on the left vertical axis. Superimposed on the incidence trends is the change in the number of seated diners from online, phone, and walk-in reservations, computed as a percentage of the corresponding number of diners one year earlier. The data, indicated by the connected dark-red line segments, are from Fort Lauderdale restaurants in the OpenTable network (OpenTable 2020). The negative numbers, shown on the right vertical axis, represent percentage declines in restaurant seating.

**Figure 4.**
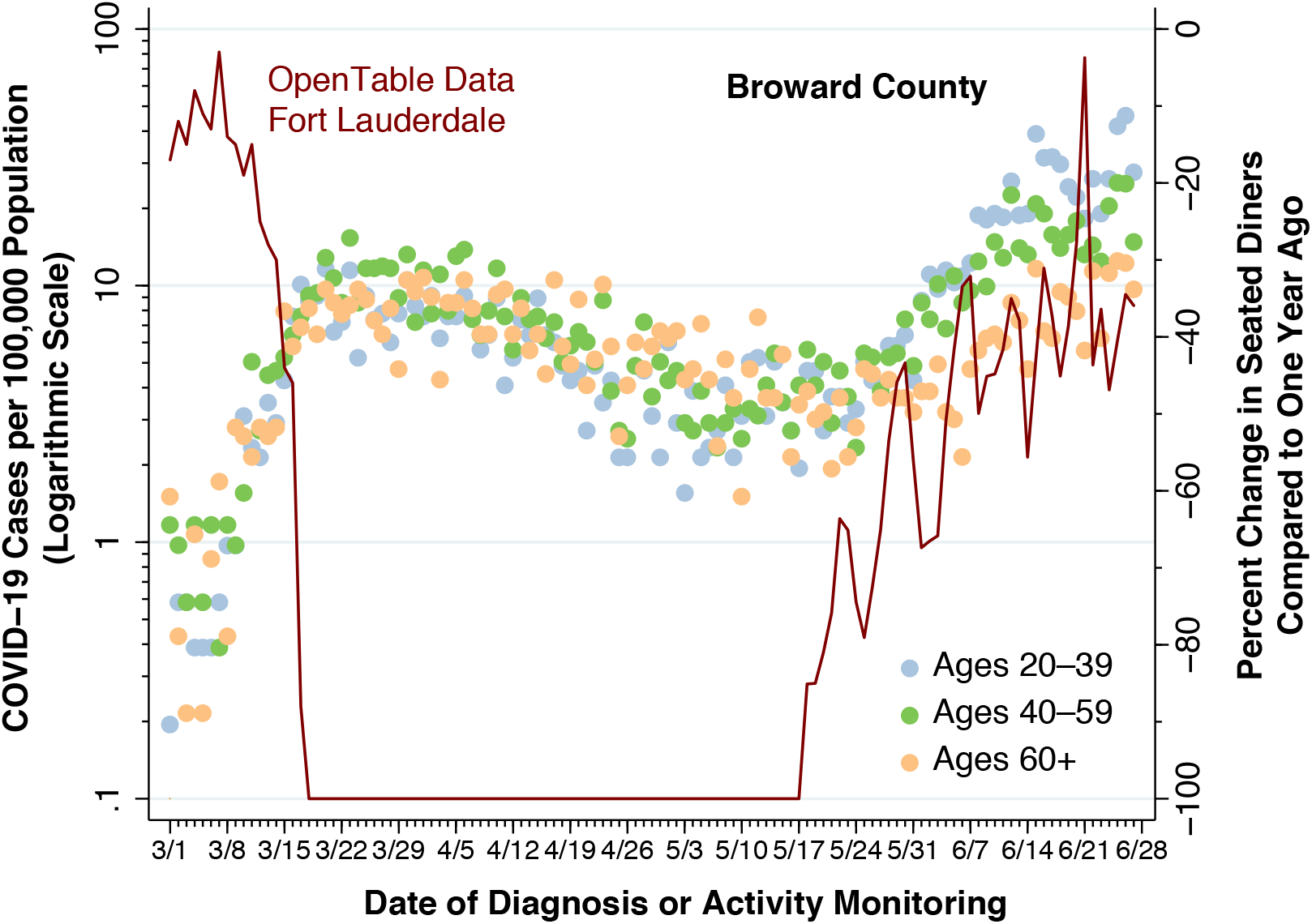
COVID-19 Case Incidence for Three Age Groups, Broward County, and Percentage Change in Seated Restaurant Diners, Fort Lauderdale Restaurants, March 1 – June 27, 2020

The data in Figure 4 show prompt, full compliance with Executive Order Number 20-68, effective March 17, as well as reopening to seated diners after Executive Order Number 20-123 (Full Phase 1) went into effect on May 18.^†^ Thereafter the rise in social mobility, as reflected in the TableOne indicator, parallels the surge in COVID-19 case incidence. While data are shown here only for Broward County (including Fort Lauderdale), there were similar patterns for Miami-Dade County (including Miami and Miami Beach), Hillsborough County (including Tampa), Orange County (including Orlando), and Collier County (including Naples).

Figure 5 shows the same data on the daily incidence of COVID-19 cases in the three age groups in Broward County, just as in Figure 4. By contrast, the superimposed data series shows the percentage change in daily visits to retail stores and recreational activity, as reported by Google’s Community Mobility Reports for Broward County in its entirety. The change is shown as a percentage of the baseline level of activity, which is calculated in relation to the median value for the 5-week period from January 3 – February 6, 2020 (Google 2020). As in Figure 4, negative numbers, shown on the right vertical axis, represent percentage declines in social activity.

**Figure 5.**
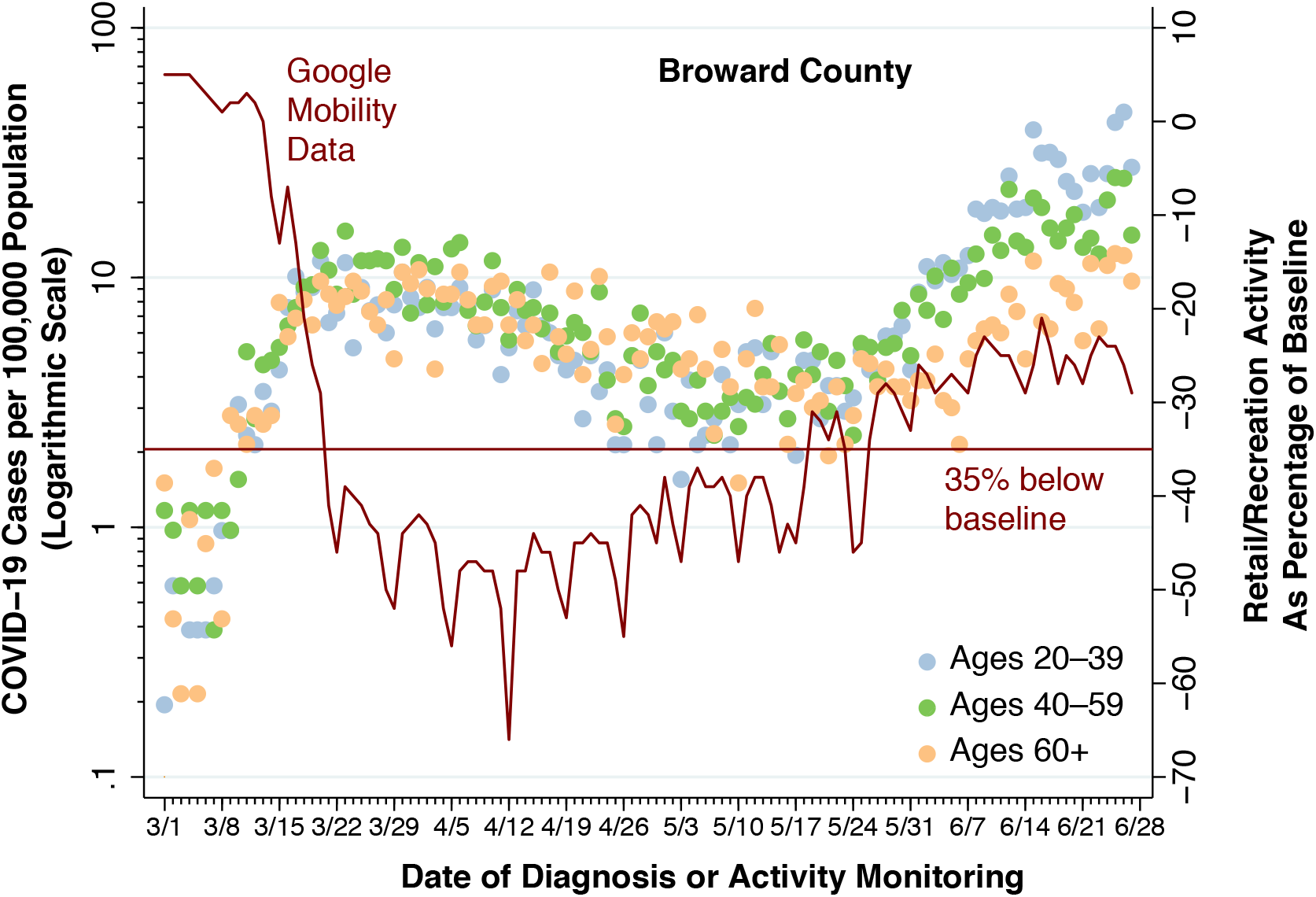
COVID-19 Case Incidence for Three Age Groups and Percentage Change in Visits to Retail Stores and Recreational Activity, Broward County, March 1 – June 27, 2020

Figure 5 likewise shows evidence of compliance with Executive Order Number 20-68, effective March 17. In contrast to Figure 4, however, the relationship between COVID-19 incidence and the Google social-mobility indicator appears to follow a threshold relationship, highlighted by the horizontal line identified as “35% below baseline.” The incidence curve decelerates when the reduction in activity exceeds 35 percent and accelerates when the reduction in activity rises to within 35 percent of baseline. The pattern observed in Figure 5 was likewise seen in the other populous counties.

### Trends in Total Tests and Positive Tests

We next inquire whether the increases in COVID-19 incidence observed from mid-May onward in Figures 2 and 3 could have been at least partly due to more liberalized testing. At the start of the epidemic in the United States, many jurisdictions initially restricted COVID-19 testing to those individuals with more severe symptoms (Harris 2020c). These restrictions were likely motivated by the scarcity of testing materials and required protective personal equipment. As testing criteria were liberalized – that is, as supply constraints were relaxed – more people with less severe symptoms would thus be expected to test positive.

To address this potential explanation, Figure 6 shows the trends in the total number of test results and the number of positive tests reported in Florida on a daily basis from March 29 through the June 27 closing date. The data for the figure, which is based upon testing for the entire state, were derived from the COVID Tracking Project (COVID Tracking Project 2020).

**Figure 6.**
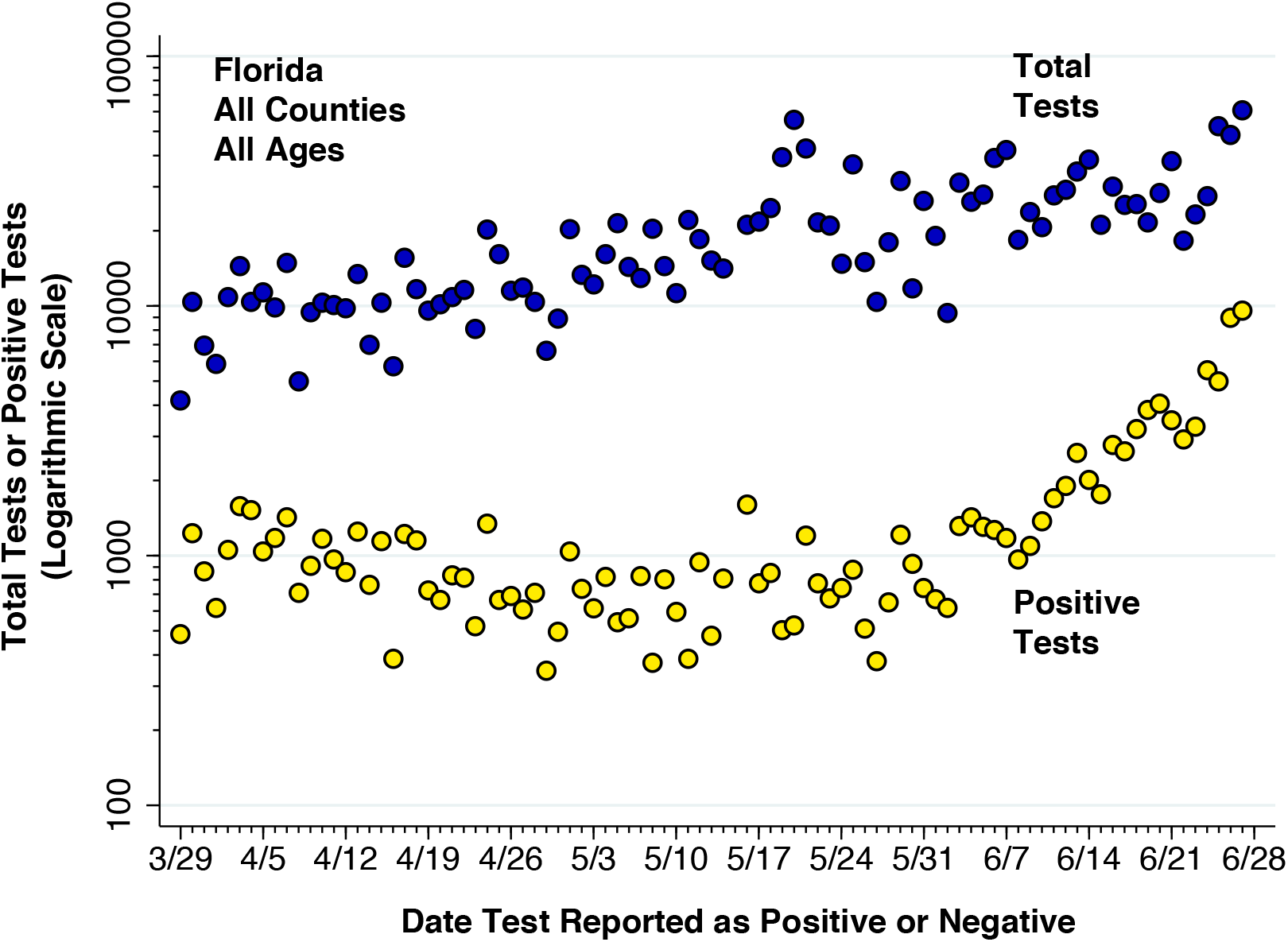
Total Tests and Positive Tests for COVID-19 Infection, State of Florida, March 29 through June 27, 2020

The dark blue datapoints represent the numbers of test results – whether positive or negative – reported each day, while the yellow datapoints represent only the numbers of positive tests reported on the same day. Thus, each test was assigned to the date it was read as positive or negative, and not necessarily to the date it was performed. The left-hand axis is shown on a logarithmic scale in order to compare proportional changes in the two data series.

Figure 6 shows that the trend in positive tests did not parallel the temporal pattern of total tests. The opening of Full Phase 1 was accompanied by a rapid expansion of mobile, walk-up and drive-thru testing (Florida Department of Public Health 2020c), with statewide tests jumping to almost 55,000 on May 20 and. Thereafter, the median number of tests was 26,380 per day (interquartile range 20,710– 37,000), peaking at 60,640 on June 27. By contrast, total positive tests initially fell as total testing rose. By the week of May 10, approximately 5.2 percent of tests were read as positive (median 5.19%, interquartile range 2.81–6.19%). Thereafter, positive tests rose much faster than total tests. By the final week of our sample, the positive test rate had increased to 15.8 percent (median 15.80%, interquartile range 9.57–18.54%).

Figure 7 shows the same lack of parallelism between total tests and positive tests for Broward County. The data were derived from the daily county reports of the Florida Department of Public Health (Florida Department of Public Health 2020a).

**Figure 7.**
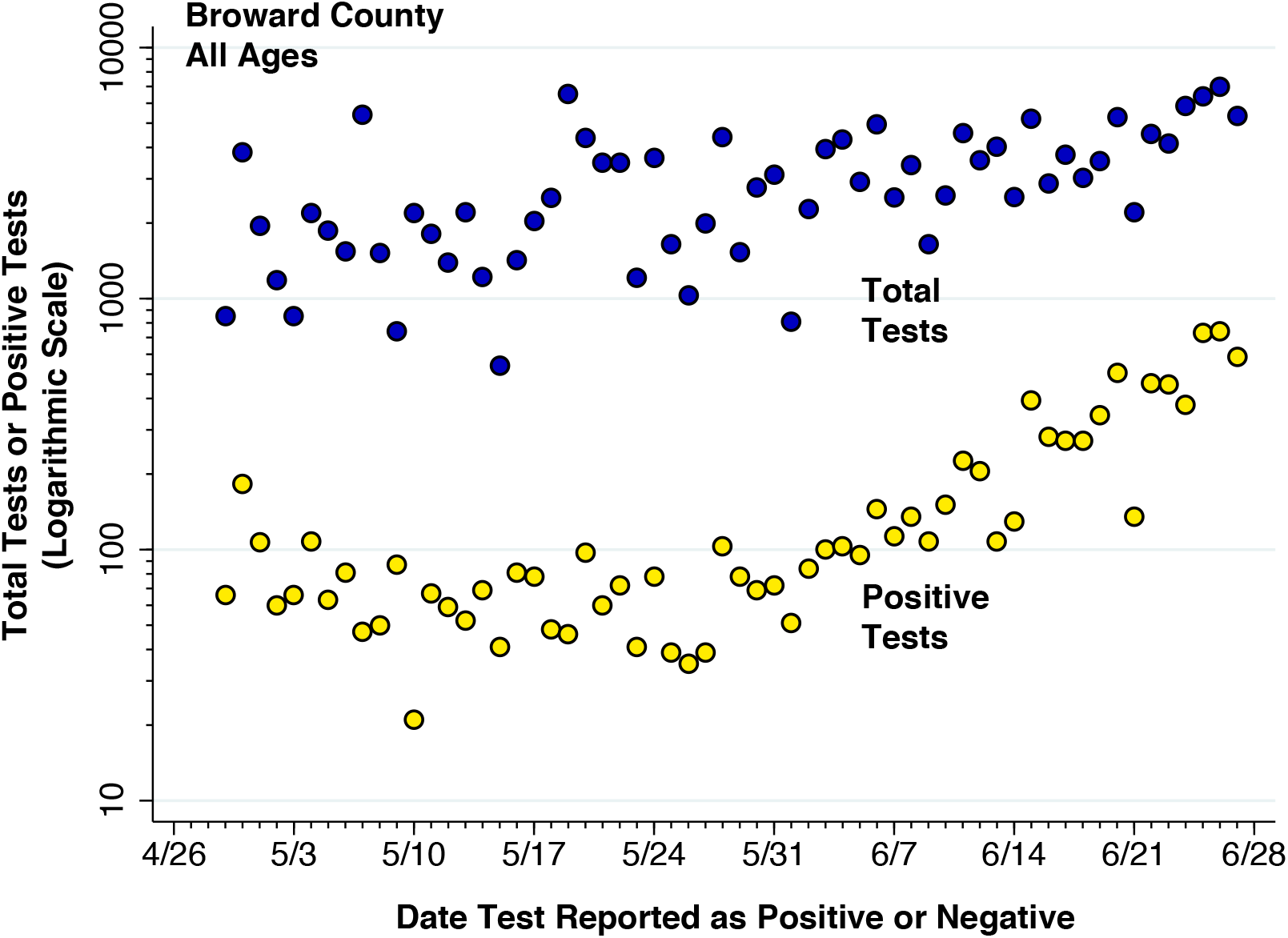
Total Tests and Positive Tests for COVID-19 Infection, Broward County, April 29 through June 27, 2020

While there is greater sampling variation, the data in Figure 7 nonetheless show an increase in total testing soon after the effective date of Full Phase 1. Positive tests, however, only gradually increased during the week of May 24. Thereafter, the increase in positive tests has substantially outstripped the change in total tests.

### Trends in Hospitalizations Among Older Persons with COVID-19

We next inquire whether recently diagnosed cases have been less severe. To that end, it would seem appropriate to examine hospitalization rates as an indicator of disease burden (Harris 2020b). Unfortunately, the data on hospitalization from the Florida Department of Health are derived from tracking positively tested individuals, and not from querying hospitals. As a result, there have been substantial delays in ascertaining recent hospitalization rates.

Figure 8 shows the trends in hospitalization status of older residents of Broward County, aged 60 years or more, who were diagnosed with COVID-19 from March 29 onward. Broward County was notable for its near-complete tracking of test-positive individuals through the end of the month of May.

**Figure 8.**
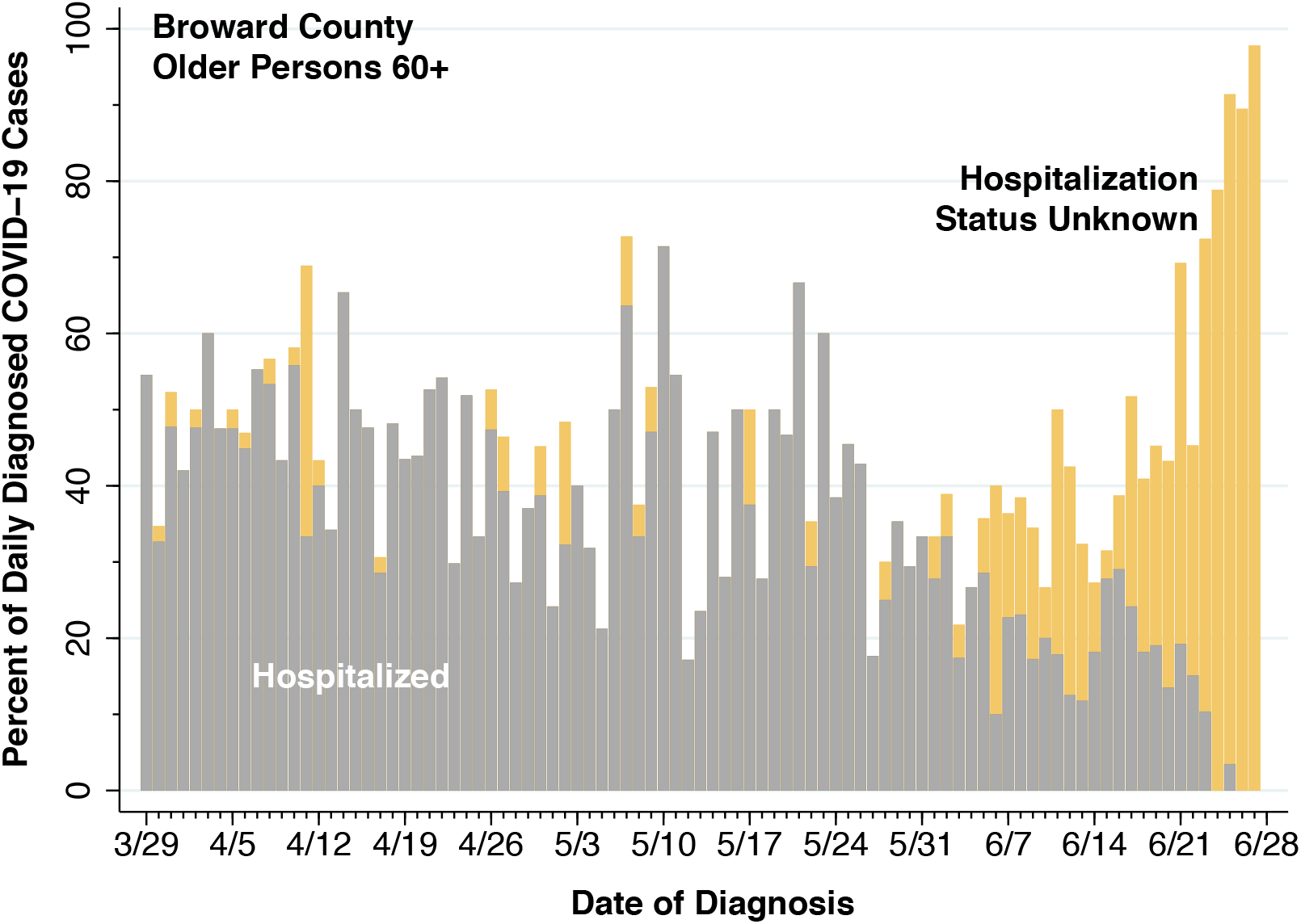
Proportions of COVID-19 Cases by Hospitalization Status, Older Persons Aged 60 Years or More Residing in Broward County, March 29 – June 27, 2020

While the gray bars in Figure 8 appear to indicate a significant decline in hospitalization rates, the mango-colored bars indicate that a growing proportion of cases has as-yet unknown hospitalization status. A similarly high proportion of recently diagnosed cases with unknown hospitalization status was seen in other populous counties. (Results not shown.) To address this data limitation, Figure 9 shows the hospitalization rates among only those older persons residing in Broward County with known hospitalization status.

Figure 9 indicates that, at least through the third week in May, there was a general downward trend in the hospitalization rate of older persons diagnosed with COVID-19 in Broward County. During the first three weeks of June, however, the hospitalization rate has been stable at about one in four infected individuals (median 26.3%, interquartile range 19.6–30.1%). At the same time, as shown in Figure 5, the incidence rate of new COVID-19 diagnoses among older persons in Broward County had increased by three-fold.

**Figure 9.**
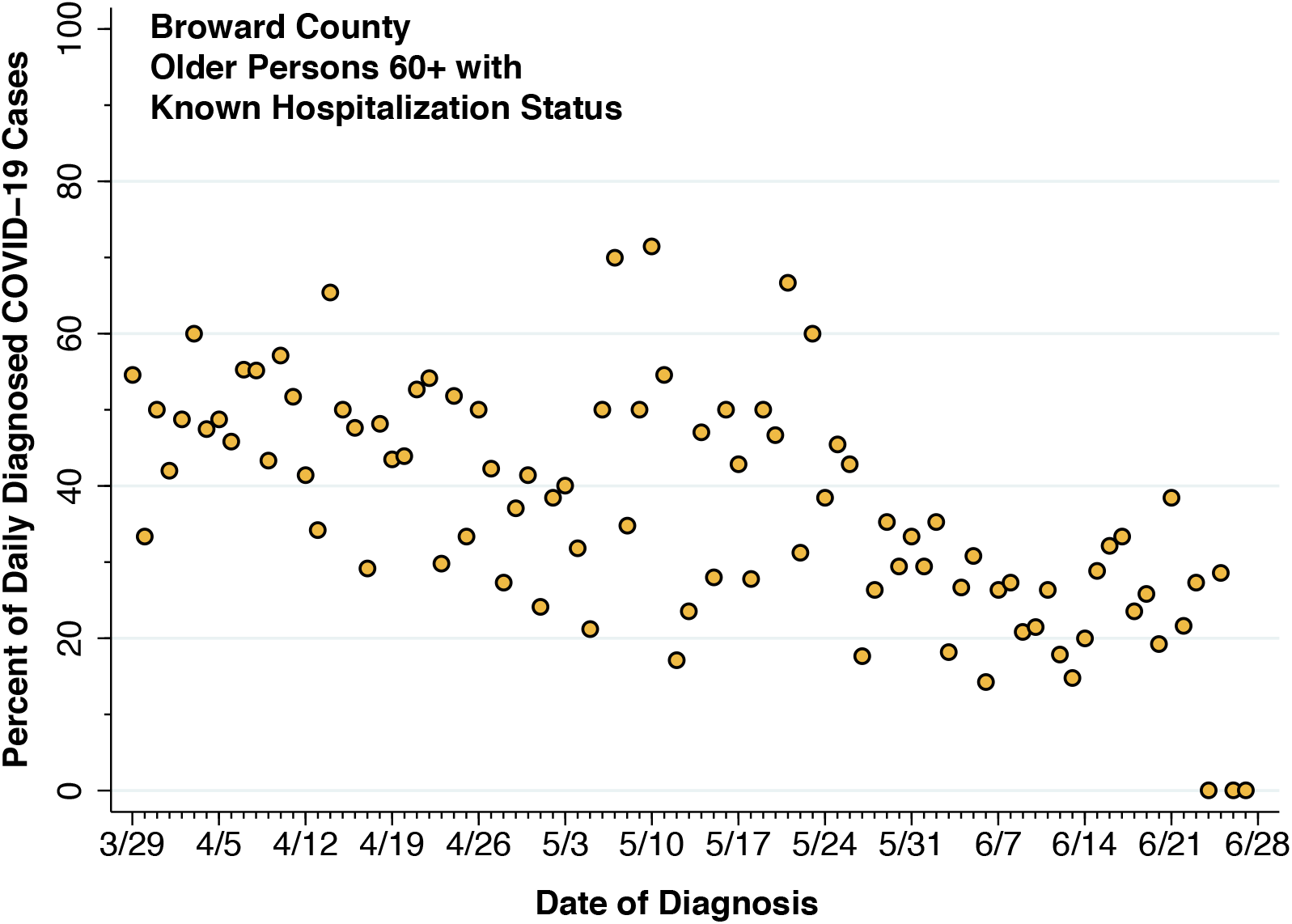
Proportions Hospitalized Among of COVID-19 Cases with Known Hospitalization Status, Older Persons Aged 60 Years or More Residing in Broward County, March 29 – June 27, 2020

### Two Age-Group Analysis

In what follows, we combine the two youngest age groups into a single age group of individuals 20–59 years of age, retaining the older group aged 60 years or more. Figure 10 plots the daily incidence of new COVID-19 diagnoses for the two broader age groups in Hillsborough County, which includes the city of Tampa. As in Figures 2, 3 and 5, we see the rise in COVID-19 diagnoses in both broad age groups, beginning soon after Full Phase 1 went into effect.^‡^

**Figure 10.**
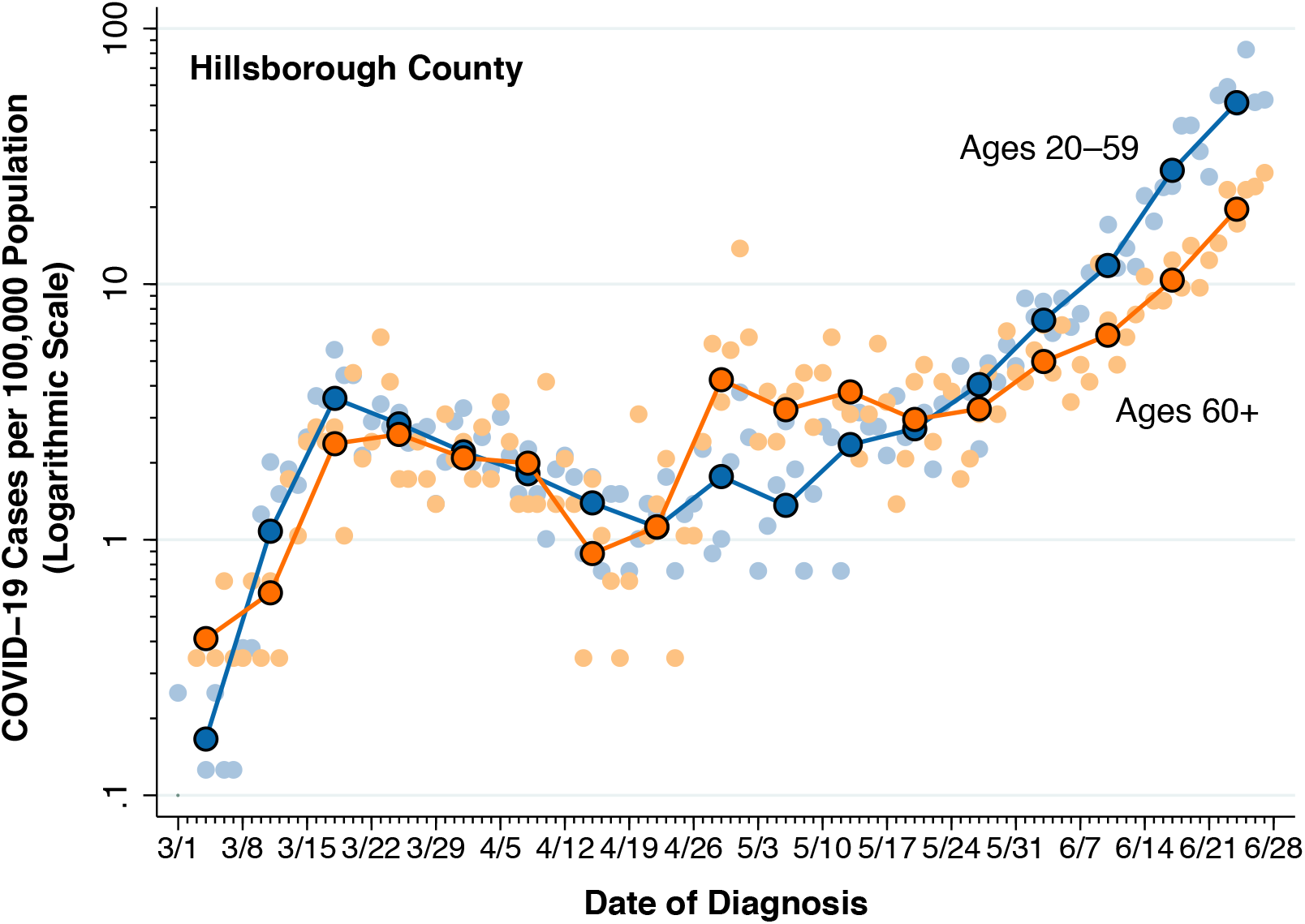
Daily Incidence of New COVID-19 Cases per 100,000 Population by Combined Age Group in Hillsborough County

Appendix 1 shows the corresponding plots for four other counties: Miami-Dade (including the city of Miami), Palm Beach (including the city of West Palm Beach), Pinellas (including the cities of Clearwater, Largo and St. Petersburg), and Volusia (including the city of Daytona Beach).

### Consistency of Trends in Incidence Across Populous Florida Counties

For each of the two combined age groups (20–59 years and 60+ years) and for each of the 16 populous counties, we used Poisson regression to estimate the daily percentage rate of increase of COVID-19 cases during Full Phase 1 from May 18 through our closing date June 17. Figure 11 plots the daily rate of increase among persons 60 years or more versus the corresponding daily rate of increase among persons 20–59 years. The size of each point is proportional to the total number of adult COVID-19 cases in each county during the Full Phase 1 period.

**Figure 11.**
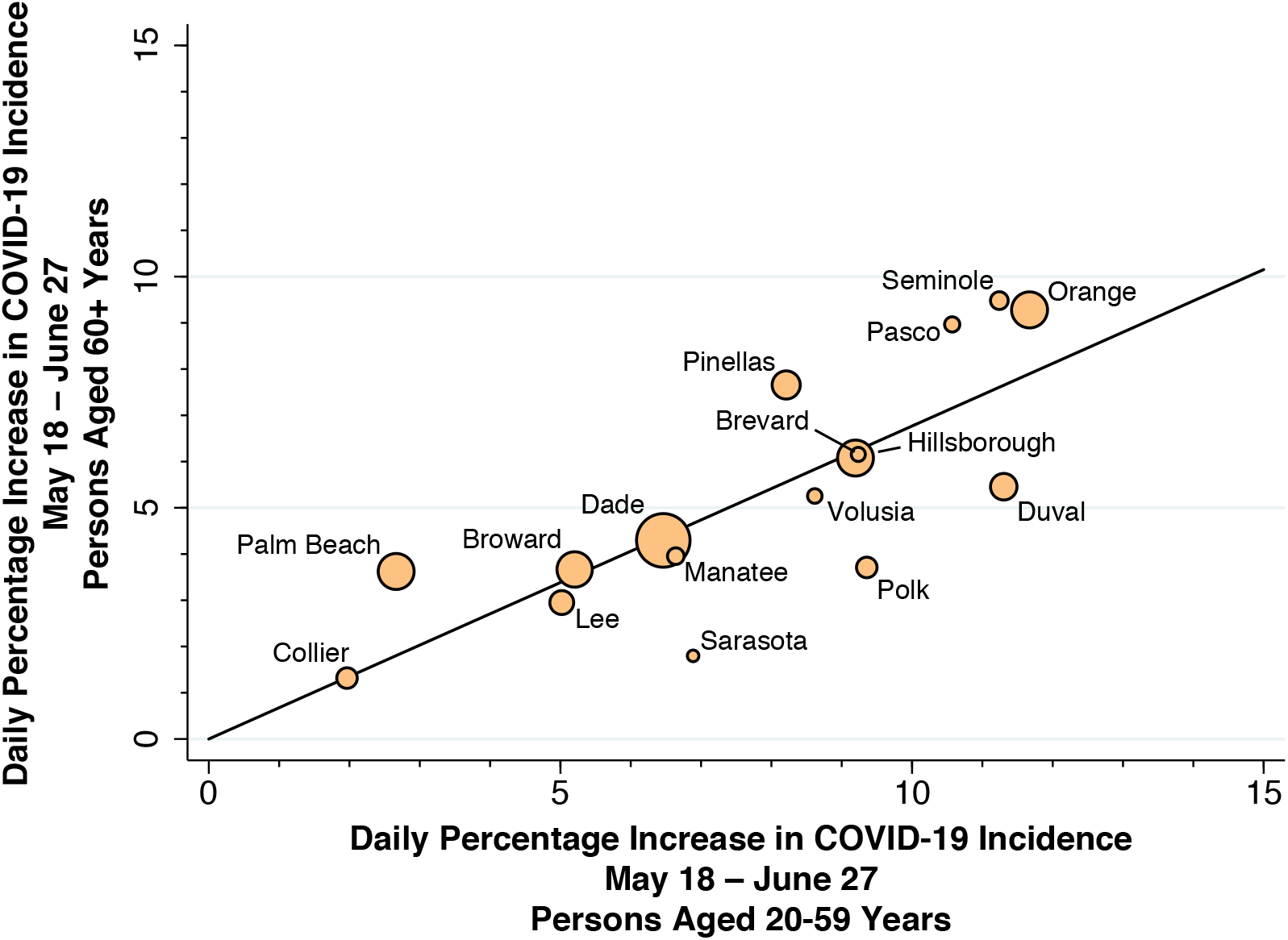
Daily Percentage Increase in COVID-19 Cases During Full Phase 1 for Persons Aged 60 Years or More Versus the Corresponding Daily Percentage Increase for Persons 20–59 Years in Each of the 16 Most Populous Counties in Florida

The plot in Figure 11 shows a consistent monotonic relationship across counties between the COVID-19 growth rates of younger and older adults during the Full Phase 1 reopening period. The slope of the best-fitting weighted least squares regression line, where the weights were the number of COVID-19 cases in each county, was +0.677 (standard error 0.141), while the unrestricted constant term was −0.0003 (standard error 0.012). That is, COVID-19 incidence among older adults aged 60 or more was on average growing two-thirds as rapidly as COVID-19 incidence among younger adults aged 20–60 years.

### Two-Group Heterogeneous SIR Model

Table 2 shows the estimated county-specific regression coefficients for the daily incidence of new infections in older persons, that is *y*_2_*_kt_* = *α*_21_*X*_21_*_k,t_*_−1_ + *α*_22_*X*_22_*_k,t_*_−1_ + *ε*_2_*_kt_*, where the regression model was run separately for each county *k*. Estimated coefficients significant at the 5-percent level (two-sided t-test) are shown in boldface, while coefficients significant only at the 10-percent level are shown in italics. Each county-specific regression had 41 observations.

**Table 2.** COVID-19 Incidence in Older Persons: Estimated Intragroup and Intergroup Transmission Parameters for Each of the 16 Populous Florida Counties in the Analytic Sample

| County | $\alpha_{21}$ | St. Error | $\alpha_{22}$ | St. Error |
| --- | --- | --- | --- | --- |
| Brevard | **0.061** | 0.025 | -0.190 | 0.172 |
| Broward | **0.052** | 0.025 | 0.052 | 0.092 |
| Collier | *0.033* | 0.019 | -0.118 | 0.134 |
| Dade | **0.269** | 0.102 | -0.181 | 0.199 |
| Duval | **0.057** | 0.015 | **0.201** | 0.086 |
| Hillsborough | **0.114** | 0.041 | -0.084 | 0.144 |
| Lee | **0.089** | 0.031 | -0.281 | 0.157 |
| Manatee | **0.041** | 0.016 | 0.085 | 0.100 |
| Orange | 0.086 | 0.057 | -0.010 | 0.200 |
| Palm Beach | 0.020 | 0.046 | **0.212** | 0.095 |
| Pasco | *0.172* | 0.101 | -0.245 | 0.301 |
| Pinellas | **0.155** | 0.037 | -0.196 | 0.113 |
| Polk | **0.085** | 0.029 | -0.067 | 0.129 |
| Sarasota | **0.017** | 0.006 | -0.046 | 0.069 |
| Seminole | **0.144** | 0.035 | -0.206 | 0.121 |
| Volusia | **0.145** | 0.040 | -0.224 | 0.159 |

Nearly all the county-specific regressions showed a significant estimate of the intergroup transmission parameter *α*_12_, reflecting the cross-infection of older by younger persons. At the same time, the intragroup transmission parameters *α*_22_ were in general not statistically significant. The notable exceptions to the overall pattern were Duval and Palm Beach Counties.

Table 3 shows the results of pooling the regressions for the 16 counties. In this specification, we allowed for county-specific interactions with the intergroup transmission variable *X*_1_*_j,t_*_−1_, but constrained the coefficient of the intragroup transmission variable *X*_2_*_j,t_*_−1_ to be uniform across counties. The omitted county in the list of county-specific interactions was Brevard County.

**Table 3.**
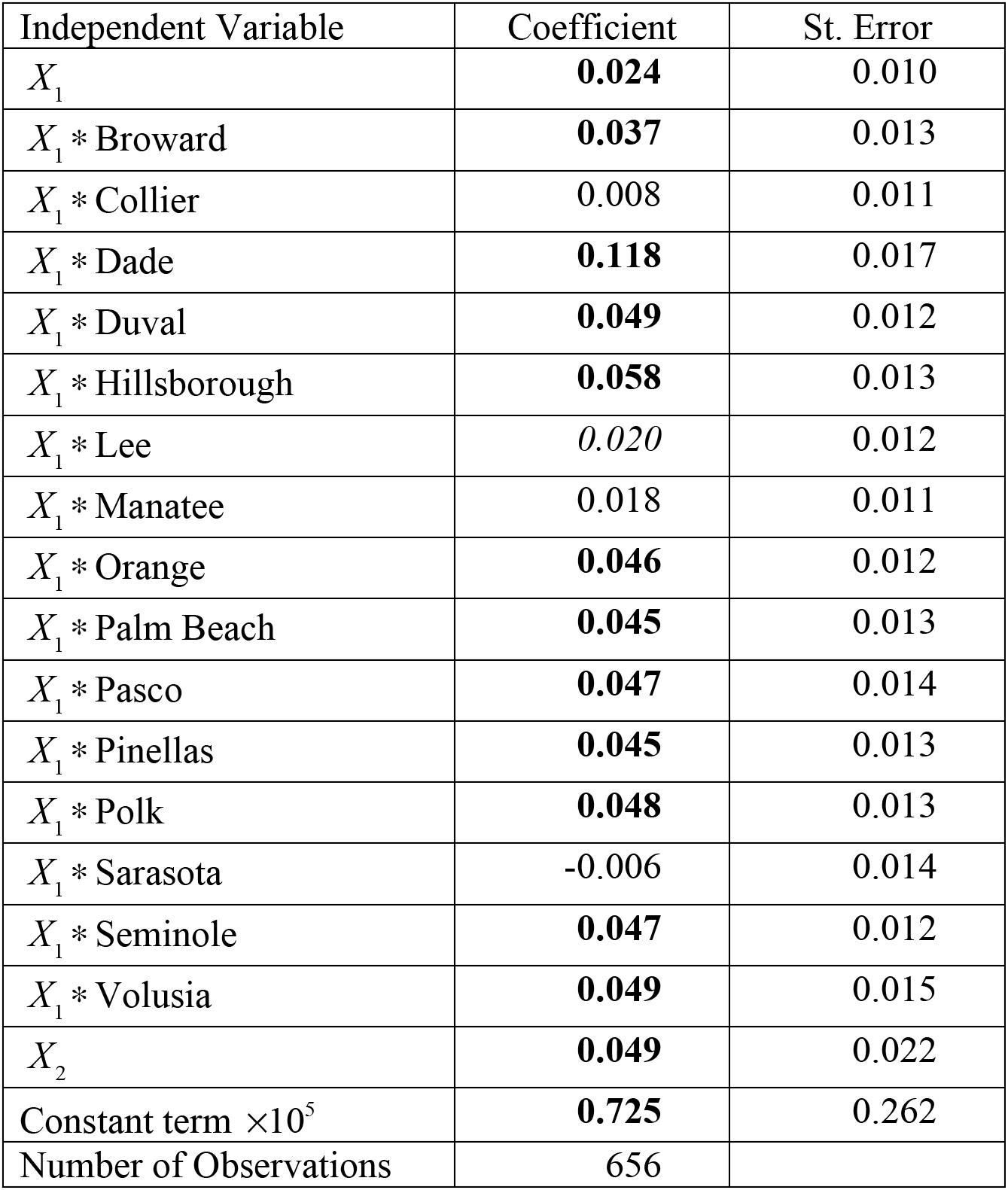
COVID-19 Incidence in Older Persons: Estimated Intragroup and Intergroup Transmission Parameters with Pooling of Florida Counties in the Analytic Sample

Again, nearly all the intergroup transmission parameter estimates were statistically significant. Pooling the data from all counties improved the precision of the intragroup parameter *α*_22_. While the constant term was estimated with precision, its estimated value of 0.725 per 100,000 population was much smaller than baseline value of 4.2 per 100,000 for all 16 counties at the start of Full Phase 1, as shown in Figure 2.

Figure 12 shows the fit of the latter model to the data on the incidence of COVID-19 infections among older persons in Hillsborough County from the Full Phase 1 reopening onward. The peach-colored datapoints are the original observations, taken from Figure 10. The connected line segments correspond to the predictions of the model.

**Figure 12.**
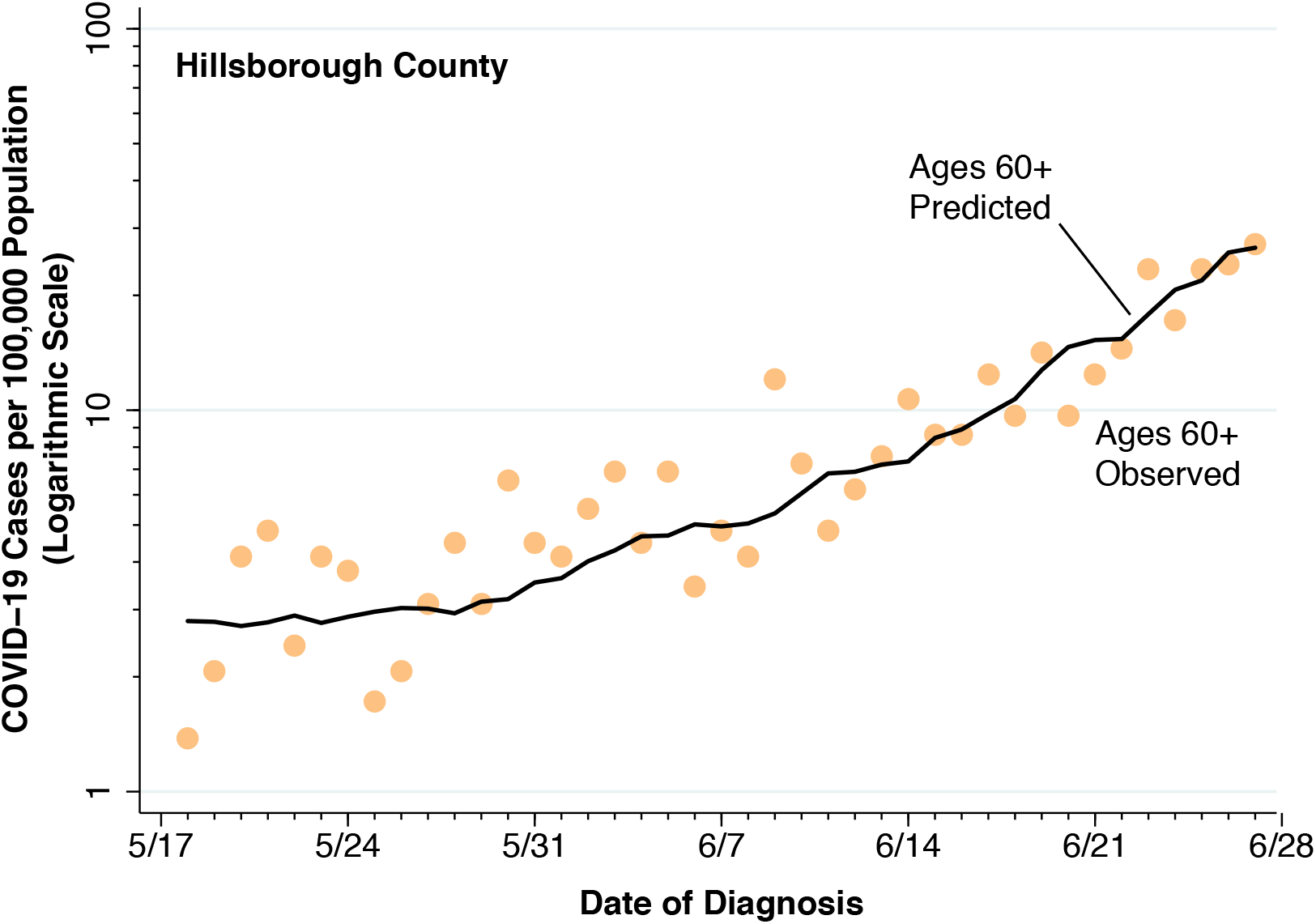
Observed and Predicted COVID-19 Incidence Among Older Persons Aged 60 or More Years in Hillsborough County Based Upon the Model Estimated in Table 3.

We also ran models of the incidence of COVID-19 infections *y*_1_*_kt_* among the younger age group. In a model analogous to that of Table 3, the estimated coefficient of *X*_11_ was 0.274 (standard error 0.015, P < 0.001). The corresponding coefficient of *X*_12_ was −0.169 (standard error 0.194, P = 0.386). (Detailed results not shown.) We also varied the common recovery parameter from *b* = l/5 to *b* = l/6. While the estimates were quantitatively different, the qualitative findings were unchanged. (Detailed results not shown.)

## Discussion

This study has a number of limitations. The basic data on confirmed COVID-19 cases was derived from a regime of partial, voluntary testing. Without universal, compulsory testing, it is difficult to draw definitive conclusions about trends in the incidence of new SARS-CoV-2 infections. Recent estimates from serologic surveys suggest that, at least in the period before the Full Phase 1 reopening, the actual incidence of infection was significantly higher (Havers et al. 2020) Nonetheless, the available evidence from this detailed study in the 16 most populous counties in Florida points to a substantial rise in case incidence in both younger and older adults after Full Phase 1 reopening went into effect on May 18.

We lack detailed data on the symptomatology and case severity of individuals voluntarily undergoing testing. Without such data, it is difficult to evaluate definitively the hypothesis that the observed rise in COVID-19 case incidence, as seen in all adult age groups in Figures 2, 3, 4, 5, 10 and Appendix 1, was due in part to liberalization of testing criteria, thus resulting in expanded testing of milder cases. Still, the patterns of total tests and positive tests seen in Figure 6 for the entire state and in Figure 7 for a specific county are inconsistent with supply constraints on testing as an important explanation for the overall rise in COVID-19 incidence. Our analysis indicates that the time path of positive tests was largely independent of the number of total tests, with positive tests rising substantially as a fraction of total tests in recent weeks.

To the contrary, Figures 4 and 5 demonstrate that indicators of social mobility, rather than measures of total testing, track the data on positive tests. These findings do not teach us that the resumption of indoor restaurant dining or visiting a retail store or an entertainment venue was the specific cause of the resurgence in COVID-19 incidence. They do, however, support the conclusion that the resurgence was the real result of changes in social mobility, and not an artifact of expanded diagnosis. Figure 5, moreover, suggests critical threshold effects in the relation between social activity and disease propagation. So long as Google mobility index remained at least 35 percent below baseline, incidence rates were declining. But when the index crossed that threshold, new COVID-19 cases surged.

We lack complete data on the hospitalization status of all persons with confirmed SARS-CoV-2 infections. The recent rapid rise in the COVID-19 caseload creates an even greater resource burden on case tracking, and thus exacerbates this problem. Without more complete data on hospitalization status, it is difficult to determine definitively whether older persons more than 60 years of age are now coming down with more or less severe cases of COVID-19. While we found evidence of a declining hospitalization rate among older persons during the earlier phases of the epidemic in Florida, we found no evidence of further changes in hospitalization rates since the recent post Full Phase 1 reopening. Hospital-based data with detailed patient information may be the best solution to this problem.

It may be difficult to determine definitively whether younger persons, having become infected as a result of increased interpersonal contact after Full Phase 1 reopening, then cross-infected older people, who remained largely at home. While more age-specific data on social mobility may be helpful, a more compelling approach will require large-scale case tracking that identifies infector-infected pairs.

Still, the evidence accumulated here is consistent with the cross-infection hypothesis. As shown in Figure 11, those counties with higher rates of increase of COVID-19 infection among young persons also had higher rates of increase among older persons. As shown in Tables 2 and 3, parameter estimates based upon a parsimonious, two-group heterogeneous SIR model indicate that the estimated cross-infection effects of young persons on older persons dominated the within-older group transmission effects. The only salient exceptions among the 16 most populous counties were the unconstrained estimates for Palm Beach County and Duval County in Table 2, where the estimated intra-group transmission among older persons was significant. These two exceptions require further study. Census data do not show these two counties to be outliers in terms of the elderly living arrangements or the proportions of elderly persons driving or employed (Florida Department of Health 2020).

There is the alternative possibility that older adults on their own engaged in enhanced social contact and, at least in principle, cross-infected their younger counterparts. Social contact matrices for the United State suggest that elderly persons have about one-third as many social contacts as younger persons (Prem, Cook, and Jit 2017). However, contact matrices capturing social interactions under normal non-epidemic conditions are unlikely to accurately represent contacts under the pressure of a persistent, life-threatening pandemic. Older persons, effectively quarantined by government order, would be more dependent on younger persons for a wide array of social needs.

Table 3 yielded an estimate of the intergroup transmission parameter *α*_22_ equal to 0.049 for older persons infecting each other. If there were no cross-infection from younger persons - that is, if *α*_21_ were zero – then the dynamic equation for the proportion *I*_2_ of infective older people would collapse to the homogenous SIR form 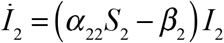. With 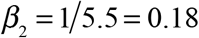 as discussed above, and with *S*_2_ ≈ 1, we would have *İ*_2_ < 0. Equivalently, the basic reproductive number would be 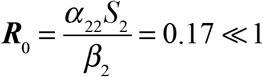 (Harris 2020a). That is, without cross-infection by younger persons, a COVID-19 epidemic solely among older, less mobile persons in Florida would be unsustainable.

An exogenous event – namely, the reopening under Executive Order Number 20-123 establishing Full Phase 1 – appears to have resulted in less strict adherence to social distancing measures by younger adults, who increasingly frequented pubs, bars, nightclubs, restaurants, beaches, retail stores, gyms, and amusement parks. These younger adults, once infected, appear to have then have cross-infected less mobile, older adults, who have largely adhered to social distancing norms.

**Appendix 1.**
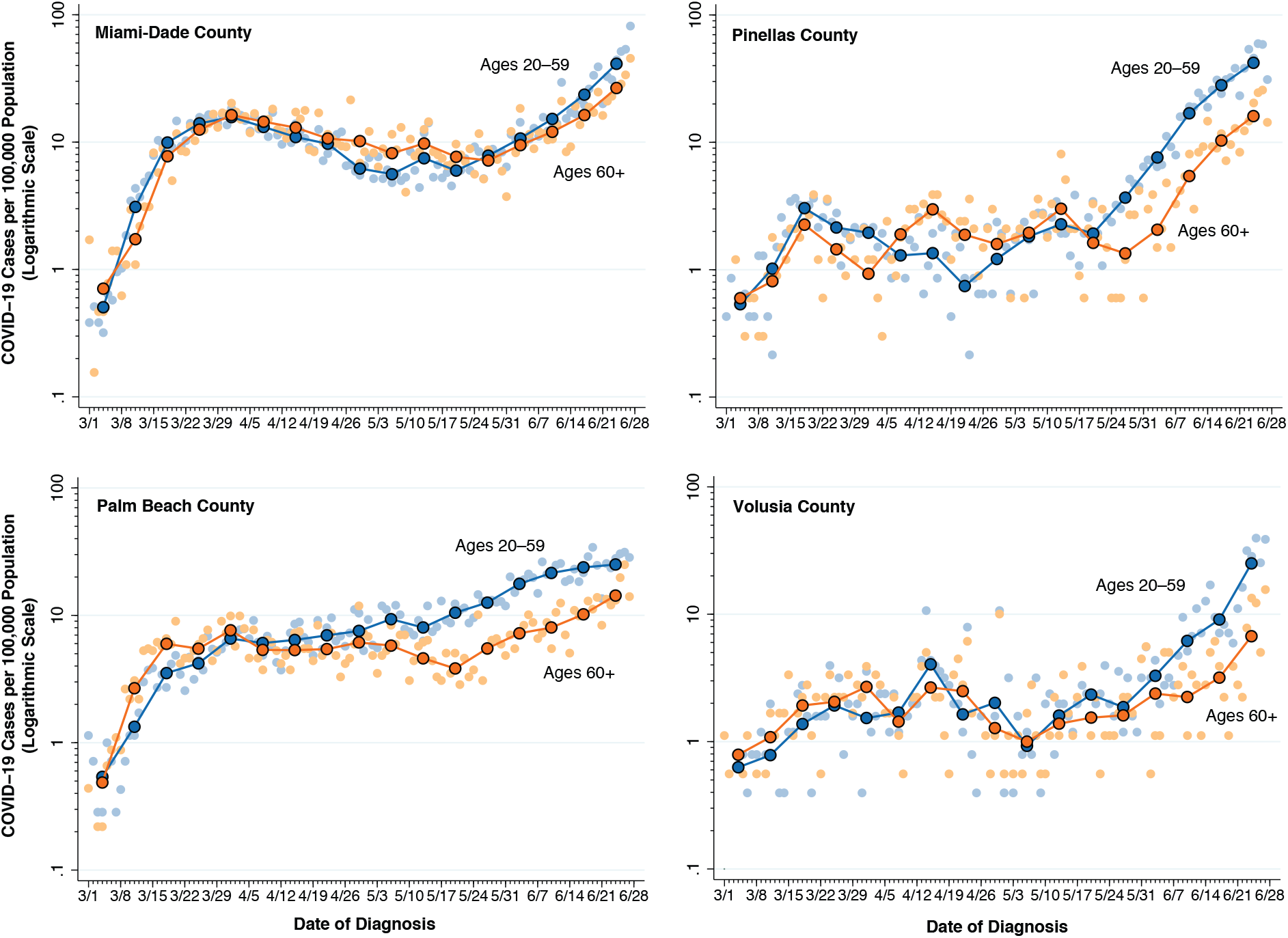
Daily Incidence of New COVID-19 Cases per 100,000 Population by Combined Age Group in Four Counties

## Data Availability

Links to the sources of all data are contained in the manuscript. All data analyses will be made available by the author.

## Footnotes

* For each calendar week, we computed the geometric mean as exp 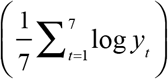, where 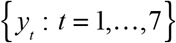 are the incidence rates for each of the seven days of that week.

† Above and beyond the governor’s statewide orders, the commissioner of Broward County had also issued emergency orders delineating guidelines, required signage, and enforcement of reopening during Full Stage 1 (Broward County 2020).

‡ Like Broward and other counties, Hillsborough has also issued its own emergency administrative orders (Hillsborough County 2020). On June 27, the closing date for this study, Mayor Jane Castor of Tampa issued an executive order requiring face coverings in any indoor location open to the public (Castor 2020).

